# Awareness, Knowledge, and Attitude toward Vasectomy among Ugandan Men: A Cross-sectional Study

**DOI:** 10.64898/2026.05.11.26352868

**Authors:** Oumo David, Chebet Fridah, Eketu Yasin, Wabwire Kaloli, Ekalu Moses

## Abstract

**Background:** Vasectomy remains one of the most underutilized contraceptive methods in Uganda, with a prevalence of only 0.2% despite its safety, effectiveness, and potential contribution to fertility reduction. Understanding the factors influencing awareness, knowledge, and attitudes toward vasectomy acceptance is crucial for developing effective promotion strategies in the Ugandan context.

**Methods:** A cross-sectional study was conducted among 617 men aged 20-60 years, selected through simple random sampling of participants attending Kapchorwa General Hospital. Data were collected using a structured questionnaire.

**Results:** Knowledge scores showed a negative association with age (β = -0.044, p < 0.001) and varied significantly by marital status, with married participants demonstrating higher knowledge than single (β = -0.624, p < 0.001) and widowed (β = -0.950, p < 0.001) individuals. Counterintuitively, higher knowledge was associated with more negative attitudes (β = -1.729, p < 0.001). Age demonstrated the strongest negative effect on attitudes (β = -0.249, p < 0.001), and 99.9% of participants believed contraception is primarily women’s responsibility. Behavioral data revealed that 75.0% desired more children, with 51.2% preferring a family size of 3-4 as the ideal.

**Conclusion:** The study shows a disconnect between knowledge, attitudes, and behaviors regarding vasectomy. While general awareness is high, deep-seated misconceptions, cultural norms around masculinity and contraceptive responsibility, and fertility preferences present significant barriers to acceptance.

## Background

Vasectomy is one of the effective, safe, and cost-efficient methods of permanent contraception, with failure rates of less than 1% and minimal associated health risks. Globally, an estimated 42-60 million men have undergone vasectomy, accounting for approximately 2.5-4.0% of contraceptive use among couples (Yang et al., 2021). However, the distribution of vasectomy acceptance shows striking geographical disparities. There are high utilization rates in developed countries like the United Kingdom (21%), the United States (12.5%), and New Zealand (22%), contrasting sharply with sub-Saharan Africa, where prevalence remains below 1% (Semu & Extension, 2023; Shongwe et al., 2019).

Despite decades of reproductive health initiatives in Africa, male-controlled methods, primarily condoms and vasectomy, remain substantially underutilized across the continent (Tumwesigye et al., 2023; Vouking et al., 2014). East Africa mirrors this pattern, with vasectomy prevalence rates of 0.1% in Kenya, 0.2% in Tanzania, and 0.3% in Rwanda, despite generally increasing contraceptive prevalence rates in the region. This underutilization persists despite vasectomy’s demonstrated advantages over female sterilization, including lower complication rates, greater cost-effectiveness, and simpler surgical procedures (Kidula et al., 2025; Mankelkl & Kinfe, 2025; Ochen & Primus, 2023).

Uganda’s reproductive health landscape is characterized by a rapidly growing population, with the current population of 45 million projected to reach 100 million by 2050. The country maintains one of the highest fertility rates globally at 5.2 children per woman, coupled with a contraceptive prevalence rate of 39% for modern methods (UBOS, 2025). Within this context, vasectomy remains exceptionally rare, utilized by only 0.2% of couples, despite its potential contribution to fertility reduction and maternal health improvement (Auma et al., 2025).

Recent global developments have renewed attention on male contraception. The growing emphasis on gender-equitable family planning, increased recognition of contraceptive method mix diversification, and emerging research on men’s reproductive health needs have created new opportunities for vasectomy promotion (Kidula et al., 2025). Simultaneously, the COVID-19 pandemic highlighted the importance of resilient health systems and long-acting methods that reduce frequent healthcare visits, potentially increasing vasectomy’s relevance in post-pandemic recovery efforts. Despite these developments, a significant evidence gap persists regarding the specific knowledge, attitude, and behavioral patterns related to vasectomy in Uganda. Most existing research has focused on general family planning or female contraceptive methods, with limited investigation of male permanent contraception (Borzuchowska et al., 2023; Sothornwit et al., 2025). Understanding the complex interplay of awareness, knowledge, and attitudes is essential for developing effective, context-appropriate vasectomy promotion strategies in Uganda.

This study therefore aimed to assess awareness, knowledge and attitude regarding vasectomy among Ugandan men, with the goal of informing evidence-based interventions to increase method acceptance and contribute to Uganda’s broader reproductive health objectives. By examining these factors within Uganda’s unique sociocultural context, this research will provide information that will support the development of targeted strategies for increasing vasectomy uptake as part of a comprehensive approach to family planning in Uganda.

## Methods

This study employed a cross-sectional survey design to assess awareness, knowledge and attitudes toward vasectomy among men in Kapchorwa, Uganda. The study population comprised adult men aged 20-60 years attending Kapchorwa General Hospital, Kapchorwa District, Uganda. The participants were selected using simple random sampling.

The sample size was determined using the formula for cross-sectional studies: n = (Z^2^ × p × q)/d^2^, where Z = 1.96 (95% confidence level), p = 0.5 (expected proportion), and d = (margin of error). This yielded a minimum sample of 587 participants. Accounting for potential non-response and the design effect, the target sample was increased by 30, to 617 participants.

### Inclusion and Exclusion Criteria

Inclusion Criteria: Biological males aged 20-60 years and willingness to provide informed consent

Exclusion Criteria: Severe cognitive impairment affecting comprehension, refusal to provide informed consent, and non-Ugandan nationals

### Data Collection Instruments and Procedures

Data were collected using a structured questionnaire developed through an extensive literature review and adapted from validated instruments used in previous reproductive health studies in sub-Saharan Africa (Ochen & Primus, 2023). The questionnaire underwent forward and backward translation into English and local languages, such as Sabiny and Kiswahili, spoken by the local community, to ensure conceptual equivalence.

The instrument comprised five sections:

Socio-demographic Characteristics: Age, education, occupation, marital status, religion, marriage type, family size, and marriage duration

Knowledge Assessment: 12 items assessing awareness and understanding of vasectomy, including permanence, safety, and effects on sexual function

Attitude Measurement: 15 items using 4-point Likert scales (Strongly Agree to Strongly Disagree) covering perceptions about male contraceptive responsibility, vasectomy safety, and gender norms

Behavioral Factors: Previous contraceptive use, fertility intentions, and ideal family size preferences

Information Sources: Channels through which participants learned about vasectomy

Data collection was conducted by trained research assistants with backgrounds in health sciences. The training included research ethics, questionnaire administration, and cultural sensitivity. Pretesting was conducted with 30 participants not included in the final sample to assess instrument clarity, timing, and cultural appropriateness.

Cronbach’s alpha for the knowledge and attitude scales was 0.78 and 0.82, respectively, indicating acceptable internal consistency.

### Variables and Measurement

Primary Outcome Variables:

Knowledge Score: Composite score (0-12) based on correct responses to knowledge items

Attitude Score: Summative score derived from attitude items, with higher scores indicating more positive attitudes

Behavioral Intentions: Willingness to undergo vasectomy and previous contraceptive practices

Independent Variables:

Demographic Factors: Age group (20-30, 31-40, 41-50, 51-60 years), education level (No education, Primary, Secondary, Tertiary), occupation (Employed, Self-employed, Unemployed)

Social Factors: Marital status, religion, marriage type, family size, marriage duration Reproductive History: Number of living children, desire for additional children

### Data Management and Analysis

Data were entered into EpiData version 4.6 and exported to R statistical software version 4.3.1 for analysis. Data cleaning included checks for completeness, consistency, and outliers. Descriptive statistics included frequencies, percentages, means, and standard deviations. Bivariate analyses employed chi-square tests for categorical variables and t-tests/ANOVA for continuous variables. Multiple Linear Regression was used to identify predictors of knowledge and attitude scores. Logistic Regression was used to assess factors associated with willingness to undergo a vasectomy. Cluster Analysis was done using K-means clustering to identify participant segments based on knowledge and attitude profiles. Model assumptions were checked, including linearity, homoscedasticity, and multicollinearity (VIF < 5 for all predictors).

Statistical significance was set at p < 0.05. Data visualization was performed using ggplot2 and patchwork packages in R.

## Results

The demographic profile of the study participants showed a predominantly married (68.2%), Protestant (60.0%) population with diverse age distribution across the four age groups (20-30: 25.0%, 31-40: 26.2%, 41-50: 25.0%, 51-60: 23.8%). The sample showed a relatively balanced distribution across key demographic variables, with traditional marriages most common (51.05%), followed by religious marriages (43.92%). Family size distribution was fairly even across categories, with 6-10 children most prevalent (26.1%), followed by 4-6 (25.1%) and 1-3 (25.0%). Educational attainment was varied, with the largest proportion having primary education (40.1%), while secondary, tertiary, and no education categories were each represented by approximately 20% of participants. Marriage duration was similarly distributed across the four categories, ranging from 23.8% to 26.2%. The details are shown in Figure 1.

**Figure 1:**
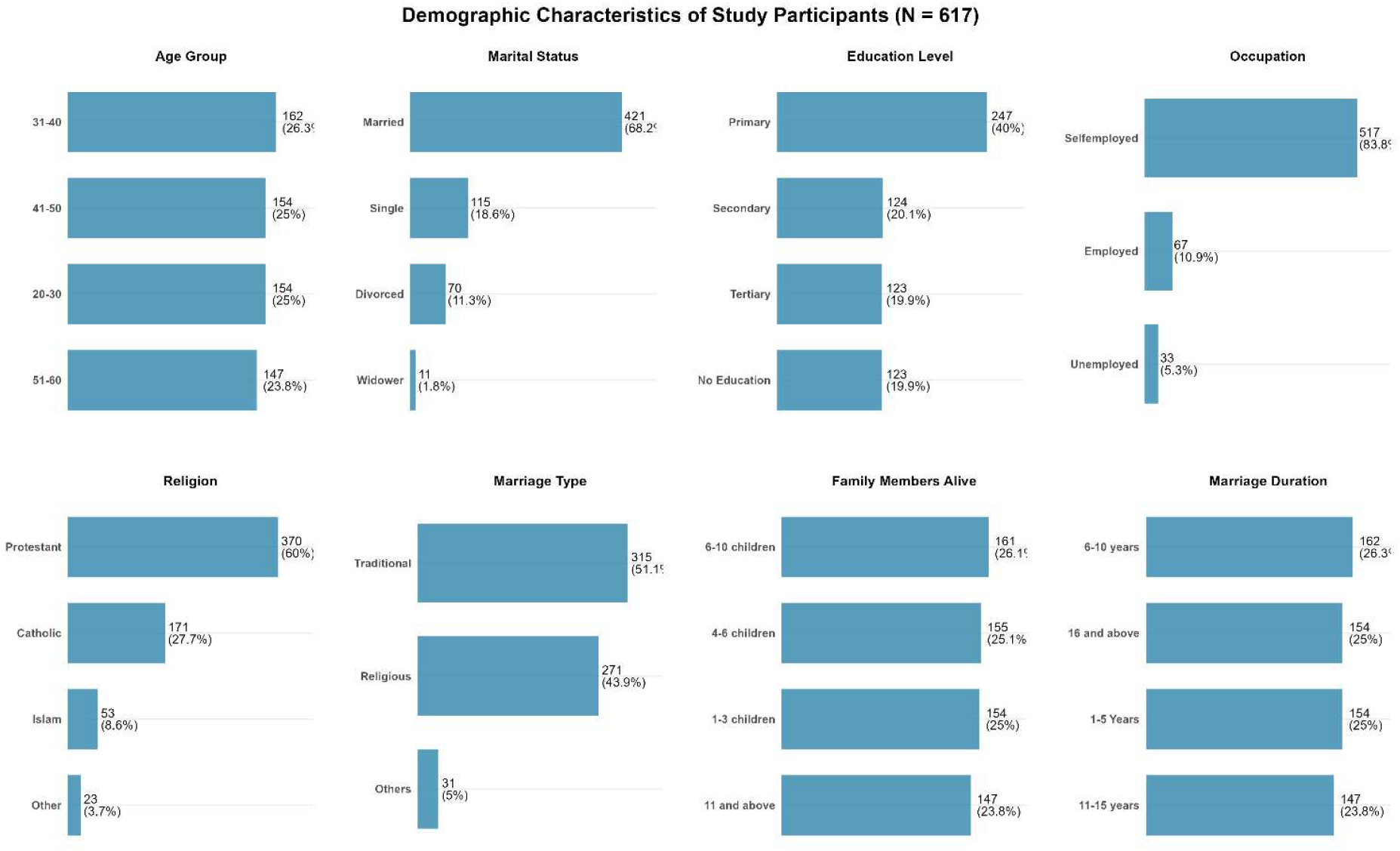
Shows the Sociodemographic characteristics of the study participants.

Knowledge levels were notably high, with 87.52% having heard of vasectomy, primarily through social media (38.57%) and health workers (37.60%). However, knowledge depth was limited, as only half of participants correctly understood that sexual function remains normal after vasectomy and that the procedure is permanent, while approximately one-quarter expressed uncertainty or held misconceptions about these key facts. Attitudes toward vasectomy were predominantly negative, with 75.04% disagreeing or strongly disagreeing that men should undergo the procedure, and 51.22% strongly disagreeing that vasectomy is better than tubectomy. Interestingly, participants overwhelmingly agreed (99.92% combined) that contraception is women’s responsibility, yet also strongly endorsed family planning benefits (75.04% agreement). Reproductive behaviors showed that 75.04% desired more children, with 51.22% preferring a family size of 3-4 as their ideal. Previous contraceptive methods were dominated by injectables (47.81%) and condoms (41.98%), indicating familiarity with family planning methods but limited adoption of permanent sterilization options. The details are shown in Table 1.

**Table 1:** Knowledge, Attitudes, and Reproductive Behaviors Regarding Vasectomy.

| Variable | Category | Count (n) | Percentage (%) |
| --- | --- | --- | --- |
| **KNOWLEDGE** |  |  |  |
| Heard of Vasectomy | No | 77 | 12.48 |
|  | Yes | 540 | 87.52 |
| Source of Vasectomy Information | Family friends | 77 | 12.48 |
|  | Health worker | 232 | 37.6 |
|  | Others | 70 | 11.35 |
|  | Social media | 238 | 38.57 |
| Normal Sexual Function | I don't know | 154 | 24.96 |
|  | No | 154 | 24.96 |
|  | Yes | 309 | 50.08 |
| Vasectomy Permanence | I don't know | 154 | 24.96 |
|  | No | 154 | 24.96 |
|  | Yes | 309 | 50.08 |
| **ATTITUDE** |  |  |  |
| Men Should Undergo Vasectomy | Agree | 154 | 24.96 |
|  | Disagree | 309 | 50.08 |
|  | Strongly disagree | 154 | 24.96 |
| Contraception is for Women | Agree | 308 | 49.92 |
|  | Strongly agree | 309 | 50.08 |
| Vasectomy is Harmful | Agree | 161 | 26.09 |
|  | Disagree | 147 | 23.82 |
|  | Strongly agree | 309 | 50.08 |
| Vasectomy is better than Tubectomy | Disagree | 301 | 48.78 |
|  | Strongly disagree | 316 | 51.22 |
| Family Planning Benefits the Family | Agree | 154 | 24.96 |
|  | Disagree | 77 | 12.48 |
|  | Strongly agree | 309 | 50.08 |
|  | Strongly disagree | 77 | 12.48 |
| **BEHAVIOR** |  |  |  |
| Need More Children | No | 154 | 24.96 |
|  | Yes | 463 | 75.04 |
| Number of Children Demanded | 1-2 children | 154 | 24.96 |
|  | 3-4 children | 316 | 51.22 |
|  | 5-6 children | 77 | 12.48 |
|  | 7 and above | 70 | 11.35 |
| Family Planning Method Used Before | COC pills | 49 | 7.94 |
|  | Injectables | 295 | 47.81 |
|  | Others | 14 | 2.27 |
|  | Use of condoms | 259 | 41.98 |

**Table 2:**
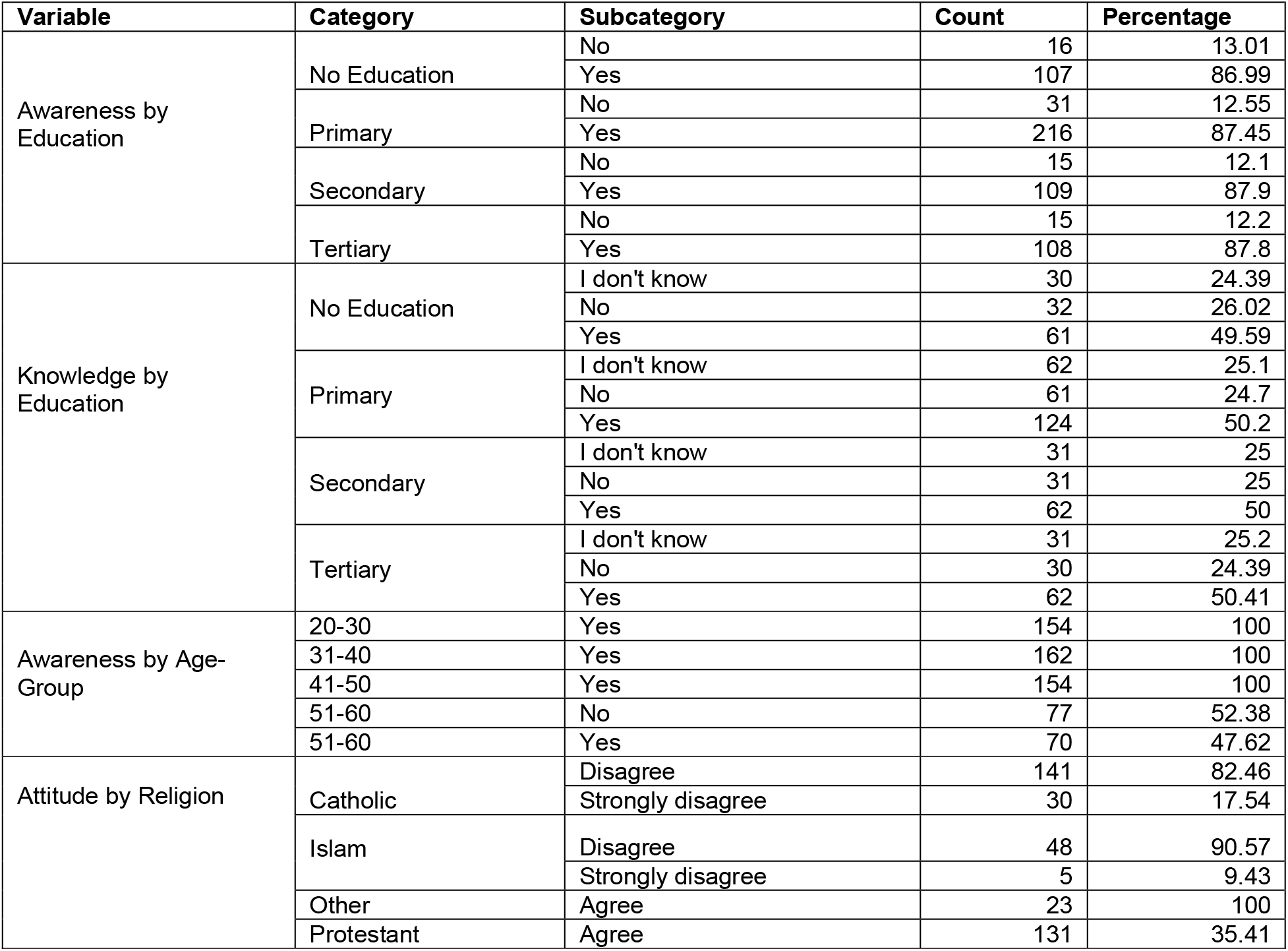
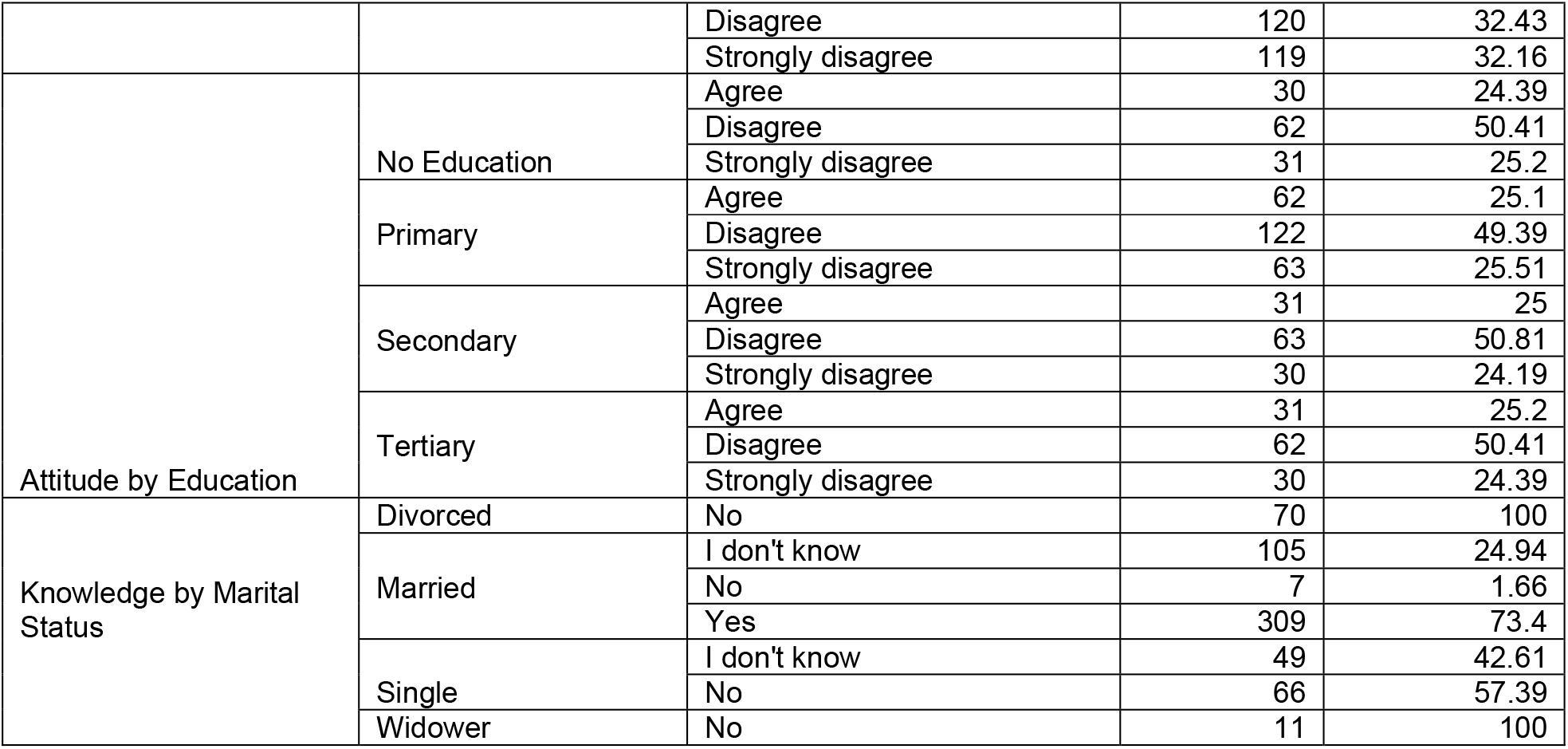
Cross-tabulation of Vasectomy Knowledge and Attitudes by Demographic Characteristics.

Universal awareness (100%) was observed among younger age groups (20-50 years), but dropped dramatically to 47.62% among those aged 51-60. Educational attainment had a minimal impact on awareness (87-88% across all education levels), but significantly influenced knowledge depth, with only about half of participants across all education levels correctly understanding vasectomy facts. Marital status emerged as a crucial factor, with married participants demonstrating substantially higher knowledge (73.40% correct) compared to single (0% correct) and divorced/widowed participants (0% correct). Occupational differences were notable: unemployed individuals showed lower awareness (84.85%) and more negative attitudes. Religious affiliation revealed stark contrasts, with Catholic (82.46%) and Islamic (90.57%) participants predominantly disagreeing about men undergoing vasectomy, while all participants from other religions supported it.

The knowledge model demonstrated a good predictive power, explaining 73.9% of the variance in vasectomy knowledge scores. Age emerged as the strongest negative predictor, with knowledge decreasing by 0.044 points per year of age. Marital status shows dramatic effects, with married individuals having significantly higher knowledge compared to single (-0.624) and widowed (-0.950) participants. Religious affiliation also significantly influenced knowledge, with Islamic participants showing higher knowledge than the Catholic reference group, while Protestant and Other religious affiliations demonstrate lower knowledge levels. Education level surprisingly showed no significant effect on knowledge scores. The details are shown in Table 3.

**Table 3:** Multiple Linear Regression - Knowledge Score Predictors.

| Predictor Variable | Coefficient ($\beta$) | Standard Error | t-value | p-value |
| --- | --- | --- | --- | --- |
| (Intercept) | 3.658 | 0.184 | 19.825 | <0.001 |
| Age | -0.044 | 0.002 | -18.799 | <0.001 |
| Education Level | 0.007 | 0.023 | 0.327 | 0.744 |
| Marital Status: Married | 0.46 | 0.078 | 5.926 | <0.001 |
| Marital Status: Single | -0.624 | 0.077 | -8.056 | <0.001 |
| Marital Status: Widower | -0.95 | 0.155 | -6.112 | <0.001 |
| Religion: Islam | 0.171 | 0.076 | 2.255 | 0.025 |
| Religion: Other | -0.62 | 0.116 | -5.356 | <0.001 |
| Religion: Protestant | -0.276 | 0.049 | -5.646 | <0.001 |
| Occupation: Self-employed | -0.042 | 0.074 | -0.56 | 0.576 |
| Occupation: Unemployed | -0.192 | 0.105 | -1.818 | 0.07 |
| R-squared | 0.739 |  |  |  |
| Adjusted R-squared | 0.735 |  |  |  |
| F-statistic | 171.9 |  |  |  |
| Model p-value | <0.001 |  |  |  |

The attitude model exhibited outstanding explanatory power, accounting for 86.6% of the variance in vasectomy attitudes. Knowledge score demonstrated a strong negative relationship with attitudes (-1.729), indicating that higher knowledge is associated with more negative attitudes toward vasectomy. Age showed the strongest effect (-0.249 per year), suggesting declining acceptance with increasing age. Marital status remained significant, with single and widowed individuals showing substantially more negative attitudes compared to married participants. Unlike the knowledge model, religious affiliation and occupation showed no significant effects on attitudes, while education level again demonstrated no meaningful relationship. The details are shown in Table 4.

**Table 4:** Multiple Linear Regression - Attitude Score Predictors.

| Predictor Variable | Coefficient ($\beta$) | Standard Error | t-value | p-value |
| --- | --- | --- | --- | --- |
| (Intercept) | 11.204 | 0.412 | 27.165 | <0.001 |
| Age | -0.249 | 0.005 | -48.197 | <0.001 |
| Education Level | 0.025 | 0.039 | 0.621 | 0.535 |
| Knowledge Score | -1.729 | 0.071 | -24.449 | <0.001 |
| Marital Status: Married | 0.237 | 0.139 | 1.707 | 0.088 |
| Marital Status: Single | -1.017 | 0.142 | -7.164 | <0.001 |
| Marital Status: Widower | -1.726 | 0.279 | -6.187 | <0.001 |
| Religion: Islam | -0.029 | 0.133 | -0.216 | 0.829 |
| Religion: Other | 0.296 | 0.206 | 1.434 | 0.152 |
| Religion: Protestant | -0.02 | 0.087 | -0.231 | 0.817 |
| Occupation: Self-employed | 0.177 | 0.129 | 1.375 | 0.17 |
| Occupation: Unemployed | 0.15 | 0.184 | 0.813 | 0.417 |
| R-squared | 0.866 |  |  |  |
| Adjusted R-squared | 0.863 |  |  |  |
| F-statistic | 354.9 |  |  |  |
| Model p-value | <0.001 |  |  |  |

## Discussion

### Awareness of vasectomy among the study participants

The findings from this study reveal a complex landscape of vasectomy knowledge, characterized by high general awareness but significant gaps in comprehensive understanding. While 87.5% of participants reported having heard of vasectomy, only 50.1% correctly understood that sexual function remains normal after the procedure, and similarly, only 50.1% recognized its permanent nature. This disparity between basic awareness and detailed knowledge aligns with recent global research demonstrating that familiarity with contraceptive methods does not necessarily translate to an accurate understanding of their mechanisms, safety profiles, and implications (Donkoh et al., 2024).

The negative association between age and knowledge score (β = -0.044, p < 0.001) is particularly noteworthy and consistent with contemporary research on contraceptive knowledge across generations. Younger participants demonstrated significantly higher knowledge levels, potentially reflecting evolving educational approaches and increased access to digital health information (Sanz-Martos et al., 2020). This generational knowledge gap has important implications for public health strategies, suggesting that targeted educational interventions for older populations may be necessary to address misinformation and knowledge gaps that persist despite broader awareness campaigns (Tripathi, 2021).

Marital status emerged as a powerful predictor of knowledge, with married individuals demonstrating substantially higher knowledge scores compared to single (β = -0.624, p < 0.001) and widowed (β = -0.950, p < 0.001) participants. This finding supports the conceptual framework that reproductive health knowledge often develops within the context of intimate relationships and family planning discussions (Al Jassem et al., 2026). The significantly lower knowledge among single individuals suggests that current educational efforts may not adequately reach populations before they enter marital relationships, representing a critical missed opportunity for early intervention (Yadassa et al., 2023).

Religious affiliation demonstrated significant associations with knowledge levels, with Islamic participants showing higher knowledge than Catholic counterparts (β = 0.171, p = 0.025), while Protestant and Other religious affiliations showed lower knowledge.

These findings contribute to the growing body of literature examining how religious and cultural contexts shape reproductive health knowledge. Recent research suggests that religious community engagement can either facilitate or hinder the dissemination of health knowledge, depending on doctrinal positions and community norms regarding family planning discussions (Arousell & Carlbom, 2016).

### Knowledge about Vasectomy among Study Participants

While 87.5% of participants reported having heard of vasectomy, only 50.1% correctly understood that sexual function remains normal after the procedure, and similarly, only 50.1% recognized its permanent nature. This disparity between basic awareness and detailed knowledge aligns with recent global research demonstrating that familiarity with contraceptive methods does not necessarily translate to an accurate understanding of their mechanisms, safety profiles, and implications (Auma et al., 2025).

The strong negative association between age and knowledge score (β = -0.044, p < 0.001) is particularly noteworthy and consistent with contemporary research on contraceptive knowledge across generations. Younger participants demonstrated significantly higher knowledge levels, potentially reflecting evolving educational approaches and increased access to digital health information. This generational knowledge gap has important implications for public health strategies, suggesting that targeted educational interventions for older populations may be necessary to address misinformation and knowledge gaps that persist despite broader awareness campaigns (Papp-Zipernovszky et al., 2021).

Marital status emerged as a powerful predictor of knowledge, with married individuals demonstrating substantially higher knowledge scores compared to single (β = -0.624, p < 0.001) and widowed (β = -0.950, p < 0.001) participants. This finding supports the conceptual framework that reproductive health knowledge often develops within the context of intimate relationships and family planning discussions. The significantly lower knowledge among single individuals suggests that current educational efforts may not adequately reach populations before they enter marital relationships, representing a critical missed opportunity for early intervention (Kumar et al., 2024).

Religious affiliation demonstrated significant associations with knowledge levels, with Islamic participants showing higher knowledge than Catholic counterparts (β = 0.171, p = 0.025), while Protestant and Other religious affiliations showed lower knowledge. These findings contribute to the growing body of literature examining how religious and cultural contexts shape reproductive health knowledge. Recent research suggests that religious community engagement can either facilitate or hinder the dissemination of health knowledge, depending on doctrinal positions and community norms regarding family planning discussions (Arousell & Carlbom, 2016; Demeke et al., 2024).

### Attitudes toward Vasectomy among Study Participants

The most striking finding was the strong negative relationship between knowledge and attitudes (β = -1.729, p < 0.001), suggesting that increased understanding of vasectomy was associated with more negative perceptions. This counterintuitive relationship challenges conventional health behavior models that typically posit knowledge as a precursor to positive attitudes and adoption. This phenomenon may be explained by the knowledge-resistance paradox, in which detailed information about vasectomy may activate specific concerns or cultural barriers that override general family planning acceptance (Kidula et al., 2025; Mankelkl & Kinfe, 2025).

The strong negative association between age and attitude scores (β = -0.249, p < 0.001) is one of the strongest effects in our analysis, indicating a dramatic decline in acceptance with increasing age. This finding aligns with recent research demonstrating generational shifts in masculinity norms and contraceptive responsibility. Older men may hold more traditional views of masculinity and gendered contraceptive responsibility, viewing vasectomy as a threat to male identity or sexual performance. This age effect persists even after controlling for other demographic factors, suggesting deeply embedded sociocultural influences that evolve across generations (Auma et al., 2025; S. Newmann et al., 2023).

Marital status continues to demonstrate significant effects on attitudes, with married participants showing more positive attitudes compared to single (β = -1.017, p < 0.001) and widowed (β = -1.726, p < 0.001) individuals. This pattern suggests that relationship context shapes contraceptive attitudes, potentially through shared decision-making processes and direct experience with family planning needs. The negative attitudes among widowed individuals may reflect life-stage considerations or the perception that permanent contraception is irrelevant outside ongoing reproductive relationships (Zimmerman et al., 2021).

The negative attitudes toward vasectomy compared to tubectomy (51.22% strongly disagreeing that vasectomy is better) reflect persistent perceptions of female responsibility for fertility management. Recent research indicates that this preference stems from complex intersections of gender norms, perceived risk assessment, and notions of bodily integrity. Participants may view female sterilization as less threatening to gender identity or sexual function, despite its greater medical complexity and risk profile (S. J. Newmann et al., 2023).

## Conclusion

The findings from this study demonstrated that knowledge alone was insufficient to drive behavioral change, as evidenced by the paradoxical negative relationship between knowledge scores and positive attitudes. The persistence of deep-seated misconceptions about sexual function and procedure permanence, coupled with strong cultural norms assigning contraceptive responsibility to women, presents substantial challenges to vasectomy promotion in the Ugandan context.

The identified demographic patterns, especially the strong negative associations with age and significant variations by marital status, highlight the need for differentiated intervention strategies. Married men emerged as the most promising target population, demonstrating higher knowledge levels and more positive attitudes, suggesting that relationship context plays a crucial role in contraceptive decision-making.

The behavioral findings, showing strong preferences for additional children and female-controlled contraceptive methods, underscore the importance of timing and method choice in family planning decisions. The near-universal attribution of contraceptive responsibility to women reflects deeply embedded gender norms that require transformative approaches rather than simple information dissemination.

## Declarations

### Ethical Considerations

The study was conducted in accordance with the principles of the Declaration of Helsinki. Ethical approval was obtained from the Research Ethics Committee of Kapchorwa General Hospital, Kapchorwa district (KGHREC138|24). Written informed consent was obtained from all participants after a detailed explanation of the study purpose, procedures, risks, and benefits. Participants were informed of their right to withdraw at any time without penalty. Confidentiality was maintained through anonymization of identifiers and secure data storage. Counseling and referral services were available to participants who required reproductive health services.

## Acknowledgments

We extend our sincere gratitude to the study participants who generously shared their time and experiences, making this research possible. Their willingness to discuss sensitive reproductive health topics demonstrates their commitment to advancing family planning in Uganda.

We acknowledge the dedicated efforts of our research team and data collectors, whose professionalism and cultural sensitivity ensured the quality and ethical integrity of data collection. Special thanks go to the district health officials and local leaders who facilitated community entry and supported the implementation of this study.

## Author contributions

OD, WK, and CF conceptualized and designed the study. OD, EM, and CF trained research assistants and supervised data collection. OD and EY handled data cleaning and analysis. CF, WK, EY, and EM drafted the initial version of the manuscript. OD, EY, EM, and EM revised the manuscript together. Both authors approved the final version.

## Data availability

The data can be requested from the authors.

## References

1. Al Jassem, O., Kheir, K., Rifi, R., Fayad, F., Kassir, R., & Salameh, P. (2026). Assessment of the knowledge, attitude, and future practices toward the concept of premarital screening among unmarried Lebanese adults: A cross-sectional study. PLOS Glob Public Health, 6(2), e0006009. 10.1371/journal.pgph.0006009

2. Arousell, J., & Carlbom, A. (2016). Culture and religious beliefs in relation to reproductive health. Best Pract Res Clin Obstet Gynaecol, 32, 77–87. 10.1016/j.bpobgyn.2015.08.011

3. Auma, A. G., Madira, E., Namukwana, B., Izaruku, R., Kabunga, A., & Te, W. M. (2025). Knowledge and perceptions of men towards vasectomy among men of reproductive age in Otuke District-Uganda. Contracept Reprod Med, 10(1), 26. 10.1186/s40834-025-00341-y

4. Borzuchowska, M., Kilańska, D., Kozłowski, R., Iltchev, P., Czapla, T., Marczewska, S., & Marczak, M. (2023). The Effectiveness of Healthcare System Resilience during the COVID-19 Pandemic: A Case Study. Medicina (Kaunas), 59(5). 10.3390/medicina59050946

5. Demeke, H., Legese, N., & Nigussie, S. (2024). Modern contraceptive utilization and its associated factors in East Africa: Findings from multi-country demographic and health surveys. PLoS One, 19(1), e0297018. 10.1371/journal.pone.0297018

6. Donkoh, I. E., Okyere, J., Seidu, A. A., Ahinkorah, B. O., Aboagye, R. G., & Yaya, S. (2024). Association between knowledge and use of contraceptive among women of reproductive age in sub-Saharan Africa. Health Sci Rep, 7(5), e2028. 10.1002/hsr2.2028

7. Kidula, N., Nguyen, B. T., Habib, N., & Kiarie, J. (2025). The impact of male contraception on global sexual and reproductive health and rights. Contraception, 145, 110811. 10.1016/j.contraception.2025.110811

8. Kumar, R., Anwar, M., Naeem, N., Asim, M., Kumari, R., & Pongpanich, S. (2024). Effect of health education on knowledge, perception, and intended contraceptive use for family planning among university students in Pakistan. Sci Rep, 14(1), 28474. 10.1038/s41598-024-79550-5

9. Mankelkl, G., & Kinfe, B. (2025). Pooled prevalence of modern contraceptive utilization and its associated factors among reproductive age women in East Africa: derived from demographic and health surveys. J Health Popul Nutr, 44(1), 261. 10.1186/s41043-025-01019-6

10. Newmann, S., Zakaras, J., Rocca, C., Gorrindo, P., Ndunyu, L., Gitome, S., Bukusi, E., & Dworkin, S. (2023). Transforming masculine norms to improve men’s contraceptive acceptance: results from a pilot intervention with men in western Kenya. Sexual and Reproductive Health Matters, 31. 10.1080/26410397.2023.2170084

11. Newmann, S. J., Zakaras, J. M., Rocca, C. H., Gorrindo, P., Ndunyu, L., Gitome, S., Withers, M., Bukusi, E. A., & Dworkin, S. L. (2023). Transforming masculine norms to improve men’s contraceptive acceptance: results from a pilot intervention with men in western Kenya. Sex Reprod Health Matters, 31(1), 2170084. 10.1080/26410397.2023.2170084

12. Ochen, A. M., & Primus, C. C. (2023). Family planning uptake and its associated factors among women of reproductive age in Uganda: An insight from the Uganda Demographic and Health Survey 2016. PLOS Glob Public Health, 3(12), e0001102. 10.1371/journal.pgph.0001102

13. Papp-Zipernovszky, O., Horváth, M. D., Schulz, P. J., & Csabai, M. (2021). Generation Gaps in Digital Health Literacy and Their Impact on Health Information Seeking Behavior and Health Empowerment in Hungary. Front Public Health, 9, 635943. 10.3389/fpubh.2021.635943

14. Sanz-Martos, S., López-Medina, I., Álvarez-García, C., Clavijo Chamorro, M. Z., Ramos-Morcillo, A., Lopez, M., Fernandez Feito, A., Navarro-Prado, S., Alvarez Serrano, A., Baena-García, L., Navarro-Perán, M., & Álvarez Nieto, C. (2020). Young Nursing Student’s Knowledge and Attitudes about Contraceptive Methods. International Journal of Environmental Research and Public Health, 17, 5869. 10.3390/ijerph17165869

15. Semu, T., & Extension, K. P. (2023). Determinants Affecting the Adoption of Vasectomy as a Family Planning Method among Married Men in Kiziranfumbi Sub-County, Kikuube District. IDOSR JOURNAL OF SCIENCE AND TECHNOLOGY, 9, 38–49. 10.59298/IDOSRJST/01.4.13111

16. Shongwe, P., Ntuli, B., & Madiba, S. (2019). Assessing the Acceptability of Vasectomy as a Family Planning Option: A Qualitative Study with Men in the Kingdom of Eswatini. Int J Environ Res Public Health, 16(24). 10.3390/ijerph16245158

17. Sothornwit, J., Lumbiganon, P., Jampathong, N., Rungreangkulkij, S., Kaewjanta, N., Kim, C., Ali, M., Bahamondes, L., Cecatti, J., Zotareli, V., Soeiro, R., Fernandes, K., Rogerio, M., Haddad, S., Bento, S., Pádua, K., Munezero, A., M’Poca, C., Laporte, M., & Camacho, G. (2025). Impact of COVID-19 pandemic on family planning and sexual transmitted infection services in Thailand: results from WHO survey. Reproductive Health, 22. 10.1186/s12978-025-02092-0

18. Tripathi, N. (2021). Does family life education influence attitudes towards sexual and reproductive health matters among unmarried young women in India? PLoS One, 16(1), e0245883. 10.1371/journal.pone.0245883

19. Tumwesigye, R., Kigongo, E., Nakiganga, S., Mbyariyehe, G., Nabeshya, J., Kabunga, A., Musinguzi, M., & Migisha, R. (2023). Uptake and Associated Factors of Male Contraceptive Method Use: A Community-Based Cross-Sectional Study in Northern Uganda. Open Access J Contracept, 14, 129–137. 10.2147/oajc.S418820

20. UBOS. (2025). Trends in family planning and related indicators. Based on the Uganda Demographic and Health Survey (UDHS) 2022.

21. Vouking, M. Z., Evina, C. D., & Tadenfok, C. N. (2014). Male involvement in family planning decision making in sub-Saharan Africa-what the evidence suggests. Pan Afr Med J, 19, 349. 10.11604/pamj.2014.19.349.5090

22. Yadassa, F., Debelew, G. T., & Birhanu, Z. (2023). The Effect of Family Planning Education on Knowledge, Attitude and Practice Toward Family Planning Methods Among Married Couples in Kersa and Goma Districts of Jimma Zone, South West Ethiopia. Risk Manag Healthc Policy, 16, 2051–2062. 10.2147/rmhp.S427176

23. Yang, F., Li, J., Dong, L., Tan, K., Huang, X., Zhang, P., Liu, X., Chang, D., & Yu, X. (2021). Review of Vasectomy Complications and Safety Concerns. World J Mens Health, 39(3), 406–418. 10.5534/wjmh.200073

24. Zimmerman, L. A., Sarnak, D. O., Karp, C., Wood, S. N., Moreau, C., Kibira, S. P. S., & Makumbi, F. (2021). Family Planning Beliefs and Their Association with Contraceptive Use Dynamics: Results from a Longitudinal Study in Uganda. Stud Fam Plann, 52(3), 241–258. 10.1111/sifp.12153

